# Hospitalizations, resource use and outcomes of acute pulmonary embolism in Germany during the Covid-19 pandemic Emergence of different phenotypes of thrombotic disease?

**DOI:** 10.1101/2021.02.08.21250291

**Authors:** Daniela Husser, Sven Hohenstein, Vincent Pellissier, Sebastian König, Laura Ueberham, Gerhard Hindricks, Andreas Meier-Hellmann, Ralf Kuhlen, Andreas Bollmann, on behalf of Helios hospitals, Germany

## Abstract

**Background:** There is discussion evolving around the emergence of different phenotypes of Covid-19-associated thromboembolic disease, i.e. acute pulmonary embolism vs pulmonary thrombosis and different phenotypes of in situ thrombosis. With this study, we wish to provide hospitalization, treatment and in-hospital outcome data for pulmonary embolism during the 2020 Covid-19 pandemic and a corresponding 2016 – 2019 control period.

**Methods:** We performed a retrospective analysis of claims data of Helios hospitals in Germany. Consecutive cases with a hospital admission between January 1 and December 15, 2020 and pulmonary embolism as primary discharge diagnosis were analyzed and compared to a corresponding period covering the same weeks in 2016 – 2019.

**Results:** As previously reported for other emergent medical conditions, there was a hospitalization deficit coinciding with the 1^st^ pandemic wave. Beginning with the 12-week interval May 6 – July 28, there was a stable surplus of hospital admissions in 2020. During this surplus period (May 6 – December 15, 2020), there were 2,449 hospitalizations including 45 PCR-confirmed Covid-19 cases (1.8%) as opposed to 8,717 in 2016 – 2019 (IRR 1.12, 95% CI 1.07 – 1.18, *P*<0.01). When excluding Covid-19 cases IRR was 1.10 (95% CI 1.05 – 1.15, *P*<0.01). While overall comorbidities expressed as weighted AHRQ Elixhauser Comorbidity Index (14.1 ± 10.1 vs. 13.9 ±10.3, *P*=0.28), the presence of thrombosis (46.1 vs 45.4%, *P*=0.55) and surgery (3.8 vs. 4.3%, *P*=0.33) were comparable, coagulopathy (3.3 vs 4.5%, P=0.01) and metastatic cancer (3.0 vs 4.0%, *P*=0.03) as contributing factors were less frequently observed during the 2020 surplus. Interventional treatments (thrombolytic therapy, thrombectomy or inferior vena cava filter placement) were less frequently used (4.7 vs 6.6%, OR 0.72, 95% CI 0.58 − 0.89, *P*< 0.01). Similarly, intensive care (35.1 vs 38.8%, OR 0.83, 95% CI 0.75 − 0.92, *P*< 0.01) and mechanical ventilation utilization (7.2 vs 8.1%, OR 0.88, 95% CI 0.74 – 1.04, *P*=0.14) as well as in-hospital-mortality rates (7.8 vs 9.8%, OR 0.76, 95% CI 0.64 − 0.90, *P*< 0.01) were lower in 2020 compared with 2016 – 2019. This was associated with a shorter length of hospital stay (6.4 ±5.4 vs. 7.2 ±5.7 days, *P*< 0.01) during the 2020 surplus period.

**Conclusions:** Only a minority of cases were associated with PCR-confirmed Covid-19 but this does not rule out preceding or undetected SARS-CoV-2 infection. Although there is a shift towards milder disease course, the increased incidence of hospitalizations for pulmonary embolism requires immediate attention, close surveillance and further studies.

---

Covid-19 infections are associated with a high prevalence of venous thromboembolism, particularly pulmonary embolism. In this respect, there is discussion evolving around the emergence of different phenotypes of Covid-19-associated thromboembolic disease, i.e. acute pulmonary embolism vs pulmonary thrombosis and different phenotypes of in situ thrombosis.^1-3^

The Helios hospital group is the largest hospital network in Germany serving about 10% of the German population. We have established a continuous surveillance program to monitor and report the effects of the Covid-19 pandemic on hospital admissions, resource use and outcomes.^4-8^ With this study, we wish to complement this discussion by providing hospitalization, treatment and in-hospital outcome data for pulmonary embolism during the 2020 Covid-19 pandemic and a corresponding 2016 – 2019 control period.

We performed a retrospective analysis of claims data of Helios hospitals in Germany. Consecutive cases with a hospital admission between January 1 and December 15, 2020 were analyzed and compared to a corresponding period covering the same weeks in 2016 – 2019 (control period). Hospitalizations were selected based on the primary discharge diagnosis of pulmonary embolism (I26) according to the International Statistical Classification of Diseases and Related Health Problems [ICD-10-GM (German Modification)]. In-hospital treatments were defined according to the German procedure classification („Operationen und Prozedurenschlüssel“, OPS) for intensive care (OPS 8-980, 8-98f or duration of intensive-care stay > 0 days), mechanical ventilation (OPS 8-70x, 8-71x or duration of ventilation > 0 hours), thrombolytic therapy (OPS 8-836.78, 8-020.8), thrombectomy (OPS 5-380.42, 8-836.88) and inferior vena cava filter placement (OPS 8-839.1), and length of stay and in-hospital mortality were calculated. For the latter only completed hospitalizations were included, i.e. patients were discharged or died in hospital (exclusion of hospital transfers). Incidence rates for admissions and treatments were calculated by dividing the number of cumulative events by the number of days for each time period. Incidence-rate ratios (IRR) or odds ratios (OR) were calculated using Poisson regression to model the number of hospitalizations and logistic regression to model the proportions of treatments per period, respectively. To identify admission trends over time, rolling IRRs were calculated for 12-week intervals with 11-week overlap resulting in one IRR every week. Inferential statistics were based on generalized linear mixed models specifying hospitals as random factor. We report IRR or OR (calculated by exponentiation of the regression coefficients) together with 95% confidence intervals (CI) for the comparisons of the different periods and *P* values for the interactions. For all tests, we apply a two-tailed 5% error criterion for significance. This study was approved by the Ethics Committee at the Medical Faculty, Leipzig University (#490/20-ek). Due to the retrospective study of anonymized data, informed consent was not obtained.

Hospital admissions and new SARS-CoV-2 infections in Germany as well as rolling 12-week hospitalization IRRs are depicted in Figure 1. As previously reported for other emergent medical conditions, there was a hospitalization deficit coinciding with the 1^st^ pandemic wave. Beginning with the 12-week interval May 6 – July 28, there was a stable surplus of hospital admissions in 2020 (Figure 1B). During this surplus period (May 6 – December 15, 2020), there were 2,449 hospitalizations including 45 PCR-confirmed Covid-19 cases (1.8%) as opposed to 8,717 in 2016 – 2019 (IRR 1.12, 95% CI 1.07 – 1.18, *P*<0.01). When excluding Covid-19 cases IRR was 1.10 (95% CI 1.05 – 1.15, *P*<0.01). Patients’ age during the 2020 surplus was similar (69.0 ±15.3 vs. 68.4 ±15.4 years, *P*=0.11) when compared to a corresponding control period in 2016 – 2019, however, with more octogenarians (30.1 vs 26.5%, *P*< 0.01). There was an observable, but non-significant change in sex distribution with more males in 2020 than in 2016 – 2019 (51.2 vs. 49.1%, *P*=0.07). While overall comorbidities expressed as weighted AHRQ Elixhauser Comorbidity Index (14.1 ± 10.1 vs. 13.9 ±10.3, *P*=0.28), the presence of thrombosis (46.1 vs 45.4%, *P*=0.55) and surgery (3.8 vs. 4.3%, *P*=0.33) were comparable, coagulopathy (3.3 vs 4.5%, P=0.01) and metastatic cancer (3.0 vs 4.0%, *P*=0.03) as contributing factors were less frequently observed during the 2020 surplus. Interventional treatments (thrombolytic therapy, thrombectomy or inferior vena cava filter placement) were less frequently used (4.7 vs 6.6%, OR 0.72, 95% CI 0.58 − 0.89, *P*< 0.01) which was mainly driven by thrombolytic therapy (4.7 vs 6.4%, OR 0.74, 95% CI 0.60 − 0.91, *P*< 0.01). Similarly, intensive care (35.1 vs 38.8%, OR 0.83, 95% CI 0.75 − 0.92, *P*< 0.01) and mechanical ventilation utilization (7.2 vs 8.1%, OR 0.88, 95% CI 0.74 – 1.04, *P*=0.14) as well as in-hospital-mortality rates (7.8 vs 9.8%, OR 0.76, 95% CI 0.64 − 0.90, *P*< 0.01) were lower in 2020 compared with 2016 – 2019. This was associated with a shorter length of hospital stay (6.4 ±5.4 vs. 7.2 ±5.7 days, *P*< 0.01) during the 2020 surplus period.

**Figure 1.**
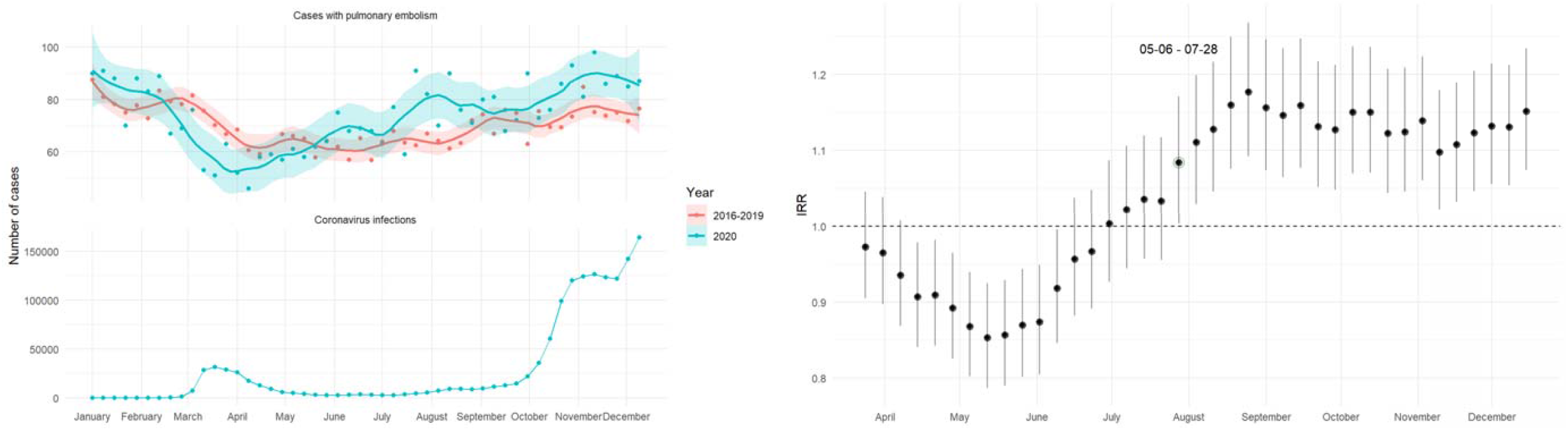
Left: total weekly hospital admissions for pulmonary embolism (upper panel) and new SARS-CoV2 infections in Germany (lower panel). Smooth curves for weekly admission rates were fitted via Locally Weighted Scatterplot Smoothing (LOESS) with a degree of smoothing of α = 0.2. Shaded areas represent 95% confidence intervals (CI). Right: rolling incidence-rate ratios (IRR) of pulmonary embolism hospitalizations in 12-week intervals with 95% CI. Please note the constant surplus in 2020 starting with the May 6 – July 28 period (IRR with circle).

By analyzing claims data of the German-wide Helios hospital network, we have identified an increase in cases with pulmonary embolism since early May 2020 compared to a corresponding period in 2016 – 2019. Interestingly, there was a slight shift in patient characteristics with respect to age and sex distribution, and less contributing factors such as coagulopathy and metastatic cancer. Although overall comorbidity burden was almost identical between cohorts, in-hospital treatments and outcomes were suggestive of less severe disease. Only a minority of cases were associated with PCR-confirmed Covid-19 but this does not rule out preceding or undetected SARS-CoV-2 infection. Taken together, those findings may support the existence of previously discussed different pulmonary embolism phenotypes associated with Covid-19.

For instance, it has been reported that Covid-19 associated pulmonary emboli are more likely to be located in the peripheral lung segments and are less extensive resulting in less frequent and severe right heart dysfunction compared to those in patients without Covid-19 pneumonia.^1^ The existence of in situ pulmonary thrombosis as one additional mechanism in patients with Covid-19 has been suggested by the absence of deep venous thrombosis, the higher incidence of pulmonary embolism but not venous thrombosis in patients with Covid-19 compared to those without Covid-19, and pathologic findings of thrombosis within the pulmonary arteries in the absence of venous thrombosis in autopsies.^1-3^

While the in-hospital course with reduced intensive care utilization, mechanical ventilation, length of stay and in-hospital mortality suggests a milder disease, the increased incidence of hospitalizations for pulmonary embolism is of special concern and could be associated with preceding Covid-19 infections.^9^ In fact, pulmonary embolism has been identified as reason for readmission after a Covid-19 hospitalization in 0.6 % of patients.^10,11^ Although this event rate seems rather low, the magnitude of Covid-19 infections worldwide may result in a substantial number of effected individuals. Alternatively or additionally, the raised awareness of respiratory symptoms that overlap between pulmonary embolism and Covid-19 such as cough or shortness of breath ^1^ may have prompted more hospital admissions. In addition, change in hospital protocols for the use of contrast-enhanced computed tomography in suspected Covid-19 cases may have detected incidental pulmonary embolism more frequently.

If the increased incidence of hospitalizations for pulmonary embolism is a signal for a rising incidence of this condition in the public, this could at least in part explain the observed excess mortality in Germany between late July and mid October 2020 not related with Covid-19 cases.^12^ This alarming finding requires immediate attention, close surveillance and further studies.

## Data Availability

The data that support the findings of this study are available on request from the corresponding author.

